# Aprepitant as a combinant with Dexamethasone reduces the inflammation via Neurokinin 1 Receptor Antagonism in severe to critical Covid-19 patients and potentiates respiratory recovery: A novel therapeutic approach

**DOI:** 10.1101/2020.08.01.20166678

**Authors:** Riffat Mehboob, Fridoon Jawad Ahmad, Ahad Qayyum, Muhammad Asim Rana, Syed Amir Gilani, Muhammad Akram Tariq, Gibran Ali, Shehla Javed Akram, Javed Akram

## Abstract

Corona virus infection is a respiratory infection, compromising the normal breathing in critical patients by damaging the lungs. The aim of this study was to evaluate the clinical outcomes of Substance P receptor Neurokinin 1 antagonist in Covid-19 patients against the usual treatments as controls.

**Methods:** It is a randomized clinical trial, open label, having two arms, one receiving normal management and care while other receiving Neurokinin-1 Receptor antagonist, Aprepitant, in addition. Dexamethasone, a corticosteroid is also administered orally to both the groups. PCR positive, hospitalized patients with more than 18 years of age, both genders, moderate to critical phase were included. 41 patients were randomly allocated in both arms, having 21 patients in group A and 20 patients in group B. Lab investigations were performed in both the groups before and after the intervention.

**Results:** Mean age of patients in group A was 50 ±18 years while 55±11 years in group B. There were 14/21 males in group A and 15/20 in group B. There were 3 critical patients in group A and 5 in group B. Biochemical and hematological parameters in both groups didn’t show much difference except the C-reactive protein reduction in the intervention group, indicative of a reduced inflammation.

**Conclusions:** The findings of this current study give a strong clue for the therapeutic potential of Aprepitant. Patients who received a combination therapy of Aprepitant and Dexamethasone showed improved clinical outcomes, laboratory findings and reduced C-reactive protein which is an inflammatory marker.

**Trial Registration:** This trial is registered in clinicaltrials.gov (NCT04468646). To Determine the Efficacy of Neurokinin 1 Receptor Antagonist as a Therapeutic Tool Against Cytokine Storm and Respiratory Failure in Covid-19 Patients

## Introduction

Novel Coronavirus infection (Covid-19) has become a pandemic in these days. It is an acute respiratory and infectious disease with no etiology and treatment known. It is continuously causing losses of precious lives and economy at a global scale on daily basis. It is need of the hour to find some treatment strategy by either preparing a vaccine or medicine or to boost the immune system. Currently, only management and prevention strategies for the spread of infection are emphasized but there is no treatment available. Different clinical trials are undergoing to evaluate the anti-viral and anti-inflammatory effects of certain drugs but still there are no concrete findings.

Substance P (SP) is a neurotransmitter and neuromodulator, released from the trigeminal nerve of brainstem as a result of nociception^1^. SP is the most common neuropeptide in the airway^2^. Coronavirus enters through the mouth, nose, eyes mainly reaching the lungs through trachea. Substance P (SP) is also released in these regions through non-neuronal cells such as immune cells. It is also released from trigeminal nerve as a result of nociception. Substance P initiates the cytokine storming via binding to its receptor neurokin-1 and many inflammatory mediators are released. If Substance P release is reduced by Neurokinin-1 receptor antagonist, may control the cytokine storming and hence the hyper-responsiveness of the respiratory tract through reduction in cytokine storming^3^.

It may be directly related to the respiratory illness as is in Covid-19 infection. It may be responsible for the increased inflammation and the signature symptoms associated with this disease. It is the main switch that need to be switched off on urgent basis by using the Neurokinin-1 Receptor (NK-1R) antagonist, Aprepitant. NK-1R is the receptor of SP and responsible for its functionality. In most complicated cases of Covid-19 infection, the clinical manifestations included fever, cough, fatigue, muscular pain, sputum production, loss of smell, shortness of breath, joint pain, sore throat, head ache, vomiting and pink eyes. In severe cases it may lead to respiratory failure, organ failure, and eventually death.

SP is present in a large amount in the CNS and the peripheral nervous system^4^. In neurons, SP is expressed in the soma. Once synthesized, SP is transported in large dense-core vesicles (LDCVs) and the peptide is released through the exocytosis of LDCVs either at axonal terminals or at the neuronal soma^5^. SP is also released by immune cells such as mast cells, macrophages, dendritic cells, T lymphocytes, neutrophils and eosinophils^6^. Substance P mediates interactions between neurons and immune cells, with nerve-derived substance P modulating immune cell proliferation rates and cytokine production^7^. Once exocytosed, SP binds to membrane-bound SP receptor expressed either on the same cell or on the neighboring cells. Unbound SP can be degraded by a cell surface metalloendopeptidase named neprilysin, and thereby suggests a shorter half-life of SP in tissues ^8^. However, SP can prolong its half-life by interacting with fibronectin ^9^. In support, SP is reported to be more stable in blood plasma ^10^. Thus, SP may form a complex with other molecules to prolong its half-life in tissue or in the blood.

Neurokinin signaling is involved in many inflammatory processes. NK-1R is a 7-transmembrane domain, G-protein coupled receptor with highest affinity for SP. Its full length form has 407 amino acids. It is present on many cell types in the body including the vascular endothelium and lymphatics, fibroblasts, white blood cells, neurons, cardioventillatory regulatory centers and phrenic nuclei controlling the diaphragm and respiration after binding to SP.^11^ It is localized in brainstem nuclei controlling the rhythmic control.^12^ SP-NK1-R complex, once formed, initiates a signaling cascade and produces inositol 1,4,5-trisphosphate (IP3) and diacylglycerol (DAG) ^13^. Importantly, the macrophages and other immune cells produce inflammatory mediators by activation of NF-kB and releases the proinflammatory cytokines.^14^ NK-1R manifests systemic inflammation as fever via cytokine-cyclooxygenase 2 prostaglandin E2 axis^15^.

Common and initial symptoms of Covid-19 infection include sore throat, loss of sense of smell and taste, pain in eyes, headache and flue^16^ and similar functions are carried out by SP once it is released from trigeminal ganglion via TrN. It provides somatosensory innervation to the orofacial region and has three branches: ophthalmic (V1), maxillary (V2) and mandibular (V3). So any alteration in its seceretion in response to viral infection may result in symptoms in orofacial region^17^. Vagal C fibers in larynx and upper airways secrete SP and cause cough. NK-1R antagonists are found to be helpful in reducing the refractory cough frequency.^18,19^ A study was conducted to assess the role of SP in clinical cough in humans. Serum SP levels were observed to be elevated in patients with persistent cough as compared to healthy controls. It was attributed to airway sensitivity in asthmatic cough.^20^ Previous studies from our own work have also revealed the involvement of SP and NK-1R in cardiorespiratory control^21^, sleep wake cycle^21^, inflammation^22^ and involvement in fatal outcomes^17^. Some of our recent unpublished work also reports similar findings of SP/NK-1R involvement in inflammation^23^, pain, pathologies, sudden deaths^24^ etc. SP and NK-1R are also found to be involved in immunology^7, 15^, lung inflammation^2, 7^, respiratory network stability ^25, 26^, heart failure ^27^and neurological and psychiatric disorders^28^. An increased expression of SP in plasma of patients with other viral infections such as HIV and HSV is also well documented^6, 29^.

SP via its tachykinin receptor, Neurokinin-1 (NK-1R) may be responsible for inflammation in Covid-19 infection. It may be potentially the main trigger which further activate the immune system. If blocked, it may stop the cascade of inflammation and hence, the infection too. The virus itself is not causing mortality but the respiratory failure, organ failure or heart attack due to cytokine storming. Hence, we propose Aprepitant in combination with Dexamethasone for the treatment of Covid-19 infection.

## Methods

### Trial Design and Participants

It is an open label, randomized clinical trial conducted at Bahria International Hospital, Lahore, to evaluate the effects of a combination therapy including Dexamethasone and Aprepitant in hospitalized patients with Covid-19. The trial was conducted in accordance with the principles of the International Conference on Harmonization Good Clinical Practice guidelines and approved by the Bahria International Hospital Ethics Committee and informed consent was obtained from the patients. 21 patients were enrolled in group A and 20 in group B (Figure 1).

**Figure 1:**
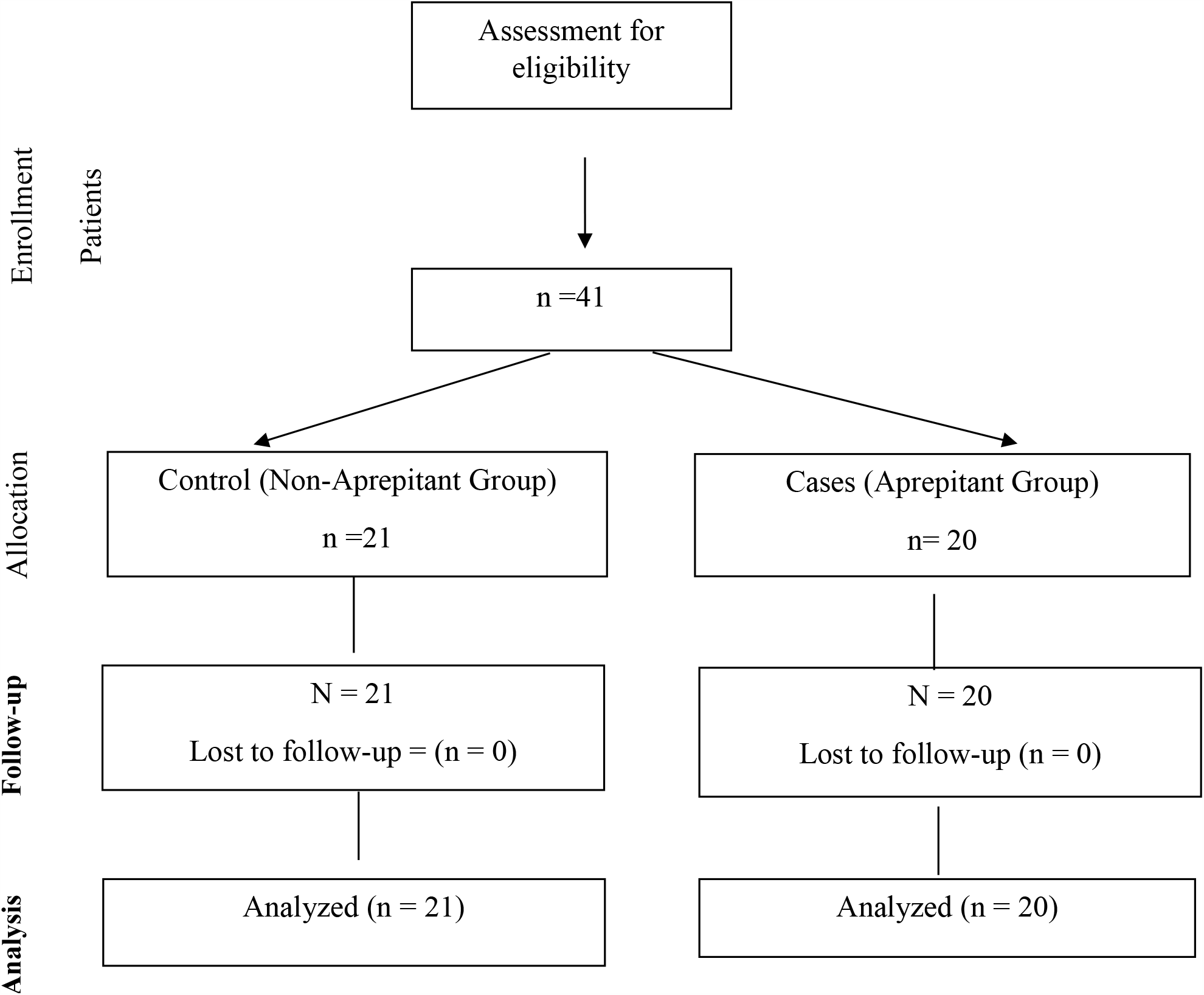
Flow diagram of the trial of two groups.

### Randomization

Case report form included demographics, major comorbidities, medication already being administered. Patients were divided randomly into two arms Control (A) and Interventioanl (B). Both the arms received routine management including Dexamethasone 20mg, but one arm received Aprepitant capsule 80mg once a day as an addition for 3-5 days depending on the condition of patient. Inclusion criteria was admitted patients above 18 years, both genders, lab confirmed Covid-19 positive based on PCR, more than 72 hours since the appearance of first symptoms. Patients with moderate to critical stages were included. Patients with respiratory diseases other than Covid-19 infection and pregnant females were excluded. Primary endpoint was total in hospital days and the duration of disease. Patients and care givers were not blinded to the allocated treatment. Lab investigations including total blood cell count, total leukocyte count, C-reactive protein, D-dimers, lactate dehydrogenase, oxygen saturation was performed on daily basis to monitor the status of patient.

## Results

### Patients

Mean age of patients in the group A (Non-aprepitant) was 50 ±18 years while 55±11 years in group B(Aprepitant)) (Table 1). There were 14 males and 7 females in A while 15 males and 5 females in B (Table1). 2 patients in group A were mild, 2moderate, 4 severe and 3 critical. In group B, 3 mild, 7 moderate, 5 severe and 5 critical. The mean WBC in group A was 13.97 ± 8.73 while in group B was 12.74 ± 5.65.(P-value >0.05). The mean PLT in group A was 293.8±68.2 while in group B was 265.8 ± 108.4 (P-value >0.05). The mean CRP in group A was 14.72±20.54 while in group B was 14.71 ± 17.47 (P-value >0.05). The mean Ferritin in group A was 512.3 ± 421.2 while in group B was525.9 ± 444.0 (P-value >0.05). The mean D-Dimers in group A was 297.23±186.28 while in group B was 486.35 ± 465.74 (P-value >0.05). The mean LDH in group A was 287.7 ± 81.8 while in group B was 418.9 ± 249.1 (P-value < 0.05) (Table 1).

**Table 1:** Baseline characteristics of demographics, clinical and laboratory findings in Non-Aprepitant and Aprepitant group

| **Demographics, clinical characteristics** |  |  |  |  |
| --- | --- | --- | --- | --- |
| **Variables** | **Total** | **Group A (Non-Aprepitant)** | **Group B (Aprepitant)** | **P value** |
| **Age** | 41 | 50 ± 18 | 55 ± 11 | 0.315 |
| **Gender** |  |  |  |  |
| **Male** | 29 | 14 | 15 | NA |
| **Female** | 12 | 7 | 5 | NA |
| **Disease Severity** |  |  |  |  |
| **Mild** | 5 | 2 | 3 | NA |
| **Moderate** | 20 | 13 | 7 | NA |
| **Severe type** | 8 | 3 | 5 | NA |
| **Critical type** | 8 | 3 | 5 | NA |
| **Comorbidities** |  |  |  |  |
| **Hypertension** | 24 | 10 | 14 |  |
| **Diabetes** | 21 | 10 | 11 |  |
| **Ischemic Heart Disease** | 13 | 5 | 8 |  |
| **Laboratory findings, Mean ± SD** |  |  |  |  |
| **WBC (1×10<sup>9</sup>/L)** | 41 | 13.97 ± 8.73 | 12.74 ± 5.65 | 0.938 |
| **PLT (1×10<sup>9</sup>/L)** | 41 | 293.8 ± 68.2 | 265.8 ± 108.4 | 0.175 |
| **CRP (mg/l)** | 41 | 14.72 ± 20.54 | 14.71 ± 17.47 | 0.657 |
| **Ferritin** | 41 | 512.3 ± 421.2 | 525.9 ± 444.0 | 0.434 |
| **D-Dimers** | 41 | 297.23 ± 186.28 | 486.35 ± 465.74 | 0.927 |
| **LDH** | 41 | 287.7 ± 81.8 | 418.9 ± 249.1 | 0.368 |
| WBC: White blood cell count; PLT:Platelet count; CRP: C-reactive protein |  |  |  |  |

Biochemical and hematological parameters of patients in group A (Table 2) and group B (Table 3) along with mean ages, disease severity status and mean values of biochemical and hematological parameters in each severity group is shown. Another remarkable finding is the platelet count which remained in normal ranges within the group B while it decreased below the normal in group A (Tables 2,3).

**Table 2:** Mean Values of Biochemical and hematological parameters in Group A Critical-CRT; Mild-MLD; Moderate-MOD; Severe-SEV

| No | Mean Age (Yrs) | Disease severity | WCC (4-11) |  | PLT (150-400) |  | CRP (>10) |  | LDH (140-280) |  | Ferritin (12-300) |  | D-Dimers (>0.5) |  |
| --- | --- | --- | --- | --- | --- | --- | --- | --- | --- | --- | --- | --- | --- | --- |
|  |  |  | Before | After | Before | After | Before | After | Before | After | Before | After | Before | After |
| 1 | 67 | CRT | 28.96 | 11.10 | 177.66 | 90.86 | 154.79 | 99.41 | 460.33 | 390.33 | 1375 | 1176 | 859.33 | 741.66 |
| 2 | 52 | MLD | 11.16 | 12.1 | 313.5 | 12.1 | 67.44 | 35.1 | 451.5 | 350 | 351.5 | 388.5 | 370 | 416 |
| 3 | 44.58 | MOD | 13.99 | 13.23 | 214.83 | 13.92 | 81.78 | 18.44 | 307.66 | 273.25 | 567.5 | 545.8 | 383.26 | 226.45 |
| 4 | 52 | SEV | 21.62 | 18.64 | 158.25 | 18.64 | 15.16 | 11.24 | 377.5 | 273.5 | 288.75 | 291.02 | 396.75 | 356.25 |

**Table 3:** Mean Values of Biochemical and hematological parameters before and after the intervention in Group B, Critical-CRT; Mild-MLD; Moderate-MOD; Severe-SEV

| No | Mean Age (Yrs) | Disease severity | WCC (4-11) |  | PLT (150-400) |  | CRP (>10) |  | LDH (140-280) |  | Ferritin (12-300) |  | D-Dimers (>0.5) |  |
| --- | --- | --- | --- | --- | --- | --- | --- | --- | --- | --- | --- | --- | --- | --- |
|  |  |  | Before | After | Before | After | Before | After | Before | After | Before | After | Before | After |
| 1 | 62.8 | CRT | 15.8 | 16.96 | 230.2 | 249.8 | 125.1 | 10.44 | 479.2 | 428.2 | 793.2 | 846 | 857.2 | 561.2 |
| 2 | 47 | MLD | 8.48 | 8.36 | 436 | 211.33 | 13.56 | 1.7 | 784.66 | 526.33 | 185.03 | 179.86 | 252.66 | 612.66 |
| 3 | 54.28 | MOD | 11.49 | 11.51 | 247.57 | 296.8 | 91.89 | 25.27 | 308.14 | 346.71 | 475.57 | 476.65 | 465.85 | 407.59 |
| 4 | 53 | SEV | 12.52 | 10.06 | 271.4 | 333.2 | 130.97 | 6.75 | 486.8 | 443.6 | 1084.4 | 535.8 | 353.6 | 226.2 |

Co-morbidities in group A and B included hypertension in 10, 14 diabetes in 10, 11 and ischemic heart disease in 05, 08 patients respectively. Among the Control group A patients, 10 were hypertensive, 10 diabetic and 5 had ischemic heart disease (Table 1). 7 patients were both hypertesive and diabetic while 4 patients were hypertensive, diabetic, had ischemic heart disease. 2 of these patients were critical, 1 was moderate and one was severe. Among the critical patients, had end stage renal stage disease and was on hemodialysis and one patient has immune thrombocytopenic purpura (ITP). 8 patients had no co-morbidity at all, most of these had mild and moderate disease severity. Mild patients had no other comorbidity, moderate had depression, asthma, obsessive compulsive disorder (OCD) and chronic obstructive pulmonary disease (COPD in one patient each. Severe patients had anti-phospholipid syndrome, acute myeloid leukemia, hodgkin’s lymphoma and obstructive sleep apnea syndrome (OSAS), in 4 patients respectively.

Among the intervention group B, 14 patients were hypertensive, 11 were diabetic and 5 had ischemic heart disease (Table 1). 11 patients were both hypertensive and diabetic. 3 patients were hypertensive, diabetic, ischemic heart disease patient as well as they had underwent Coronory Artery Bypass Grafting (CABG) previously. 2 of these patients were critical. 6 patients had no co-morbidity, while one had only migraine. One of these patients was in critical group, while, 2 in mild, one in moderate and 2 in severe disease condition group. The patient with only migraine was in Severe disease group.

Azithromycin, Penitrax, Merosol, Remdisivir, Tocilizumab were administered in both the groups as routine management and usual care, irrespective of the group allocation. Clexane was used as anticoagulant is both groups. One patient was discharged in both groups within first 5 days while one patient expired in both groups.

## Discussion

These initial findings show that a combination therapy of Dexamethasone and Aprepitant is effective in treating severe to critical Covid-19 patients. Dexamethasone is a recently discovered drug for the treatment of critically ill Covid-19 patients. It is previously being used for similar indications such as post surgery or post-chemotherapy emesis, nausea. Previously, it is in use alone as well as a combination therapy with Aprepitant. There are no major side-effects and safe for use in children as well as pregnant females. Rather it is used for lung maturation in premature labor and administered a day to week before delivery^30^. In our recent study, we have also suggested the similar role for aprepitant.^24^ It may also improve the respiratory conditions in pre-mature labour as well as Covid-19 patients. We also suggest Ondansetron, a serotonin receptor antagonist along with these two drugs. This medication is already in use for the treatment of nausea and vomiting post chemotherapy. Serotonin is also involved with the nociceptive stimuli along with Substance P in inflammation. It may also be helpful in management of critically ill patients with respiratory problems as a result of Covid-19 infection.

Although mean of all the data is not showing much significant findings but a close look at the data of individual patients reveals a marked reduction in the levels of C-reactive protein in the intervention group B as compared to the group A. It is also noteworthy, that there were more male patients in both the groups. The more critical patients were also males. There were some confounding factors as well e.g the medication that was given to both the groups including antibiotics, antico-agulants, Remdisivir and Tocilizumab, although it was given to both the groups. As the patients were randomly allocated to both the groups, coincidently, there were more aged patients in intervention group B and they had more co-morbidities. It may also affect the results.

SP secreted by the immune cells stimulates Th1 and Th17 autoreactive cells that migrate to the CNS and target nerves^31^. Hence, SP has been observed to be an important player in autoimmune responses. Expression of SP receptors, NK-1R is increased during inflammation, mainly on lymphocytes and macrophages. Hence, SP has a key role in immune system regulation by modulating the chemokines and adhesion molecules^7^. SP promotes angiogenesis by stimulating immune cells^32^. It affects the endothelial cells directly to produce nitric oxide or indirectly by interacting with mast cells and granulocytes^33^.

Neutral endopeptidase (NEP) also known as Enkephalinase is SP degrading enzyme and animal studies have reported a decreased NEP activity in trachea of rats infected with parainfluenza virus type 1, corona virus and *Mycoplasma pulmonis*. NEP depletion in respiratory tract may lead to increased vulnerability to neurogenic inflammation ny permitting larger concerntrations of SP to reach NK-1R in the respiratory tract. This reduction of NEP activity elicits the pathological responses in respiratory infections ^1^. Although SP is a neurotransmitter but its exocrine role within the respiratory tract in well known^1^. Exogenous SP is involved in the mediation of inflammation in airways^34^, causes smooth muscle contraction ^35^and mucous secretion from tracheal glands ^36^and increased vascular permeability^37^. Stimulation of sensory nerves by irritants including viruses causes bronchoconstriction^38^ and also increases the attachment of neutrophils to the endothelium of venules^39^, leading to inflammation^1^. Several aspects of SP in airways are regulated by NEP^35^. So, it is already a proven fact that respiratory tract infections potentiates the vascular permeability produced by SP due to a decrease in NEP activity in the respiratory tract. The understanding of this mechanism is crucial in treatment of respiratory infections such as Covid-19. Alterations in NEP activity may result in altered SP concentrations in airways and leading to clinical manisfestations of respiratory disease^1^.

Hence, dexamethasone, a corticosteroid may modulate immune-mediated lung injury and reduces the progression to respiratory failure and death by mainly affecting the NEP and Aprepitant may block the receptor of SP. Both dexamethasone and Aprepitant are affecting the SP, one by potentiating the SP degrading enzyme and the other by antagonizing the SP receptor, therefore, reducing the SP binding the receptor. So, the limitation of this study is small sample size as the number of patients decreased in Pakistan and we couldn’t enroll more patients. Here, we suggest, to conduct this trial in other populations were there is still surge of Covid-19 and ruling out the confounding variables. But this study gives a strong clue for the effectiveness of this treatment for severe to critical Covid-19 patients.

## Conclusions

Substance P (SP) is a possible cause of the initiation of cytokine storming developed in Covid-19 infection and we suggest Neurokinin-1 Receptor antagonist, Aprepitant, as a drug to be used for its treatment. It may help in the management of cytokine storming as well as improving the respiratory outcome in severe to critical patients. Aprepitant also affected the platelet levels and maintained within a normal range.

## Data Availability

Data will be made available on request

## Author Declarations

The trial was conducted in accordance with the principles of the International Conference on Harmonization Good Clinical Practice guidelines and approved by the Bahria International Hospital Ethic Committee.

